# Preterm birth is associated with immune dysregulation which persists in infants exposed to histologic chorioamnionitis: a descriptive study

**DOI:** 10.1101/2021.04.29.21256310

**Authors:** Gemma Sullivan, Paola Galdi, Nis Borbye-Lorenzen, David Q. Stoye, Gillian J. Lamb, Margaret J. Evans, Kristin Skogstrand, Siddharthan Chandran, James P. Boardman

## Abstract

**Objective:** To characterise the umbilical cord blood immune profile in preterm infants compared to term-born controls and the postnatal immune response following exposure to histologic chorioamnionitis (HCA) in preterm infants.

**Design:** Descriptive, observational cohort study.

**Setting:** Edinburgh, UK.

**Population:** 118 preterm infants (mean gestational age 29^+0^ weeks, range 23^+2^ to 32^+0^) and 59 term-born controls.

**Methods:** Placental histopathology was used to identify reaction patterns indicative of HCA, and a customised immunoassay of 24 inflammatory markers and trophic proteins selected to reflect the perinatal immune response was performed on umbilical cord blood in term and preterm participants and postnatal day 5 blood in the preterm group.

**Results:** The umbilical cord blood immune profile classified gestational age category with 86% accuracy (95% CI 0.78-0.92), p-value=1.242×10^−14^. Pro-inflammatory proteins IL-6, MCP-1 and CRP were elevated in the cord blood of preterm infants whilst BDNF, C3, C9, IL-18, MMP-9 and RANTES were decreased, compared to infants born at term. In preterm infants, exposure to HCA was associated with elevations in 5 immune proteins on postnatal day 5 (BDNF, C3, IL-8, MIP-1β and MMP-9) when compared to preterm infants who were not exposed.

**Conclusion:** Preterm birth is associated with a distinct immune profile in umbilical cord blood and infants exposed to HCA experience specific alterations in immune function that persist to day 5 of postnatal life.

## Introduction

Perinatal immune processes have a crucial role in neurodevelopment and early life inflammatory exposures are associated with an increased risk of neuropsychiatric disorders such as autism spectrum disorder, schizophrenia, bipolar disorder and depression ^1,2^. Preterm infants may be exposed to multiple episodes of perinatal infection/inflammation and are particularly vulnerable to brain injury resulting from a dysregulated immune response during a critical period of CNS development ^3^.

Preterm infants have a distinct immune profile in umbilical cord blood and cerebrospinal fluid that includes higher levels of pro-inflammatory cytokines and lower levels of growth factors when compared to term-born controls but there is uncertainty about the extent to which this is influenced by antenatal factors, environmental exposures and/or developmental regulation ^4,5,6^. Histologic chorioamnionitis (HCA) is strongly associated with preterm birth ^7, 8^ and increases the risk of neonatal morbidities including lung disease, intraventricular haemorrhage, sepsis and necrotising enterocolitis ^9-14^. HCA has also been implicated in the development of white matter injury, cerebral palsy and neurodevelopmental impairment ^15-18^ and this may be mediated by specific pro-inflammatory cytokines ^19^. When HCA involves a fetal inflammatory response (FIR), these risks appear to be increased further, suggesting that organ injury is mediated by a systemic fetal inflammatory response syndrome (FIRS). FIRS was initially defined using threshold values of IL-6 concentration in umbilical cord blood ^20, 21^ although subsequent studies have shown that histopathological FIR is associated with elevated concentrations of cytokines (IL-1β, IL-6 and TNF-α), chemokines (IL-8, MCP-1, MIP-1β, RANTES), matrix metalloproteinases (MMP-1 and MMP-9) and CRP^19, 22-26^. In some preterm infants, blood concentrations of inflammatory mediators remain elevated for weeks after birth ^27,28^ and may be associated with higher circulating levels of neurotrophic growth factors ^29^. However, neurotrophic capability following exposure to intrauterine inflammation is not well understood and previous study designs leave uncertainty about the role of the complement system in perinatal inflammation, which plays a critical role in the innate immune response.

Preterm infants could benefit from immunomodulatory therapies in the perinatal period but development of rational treatment strategies requires improved characterisation of the neonatal immune profile and the postnatal response to HCA. In this study, an immunoassay of 24 analytes customised to reflect the perinatal immune response was used to analyse profiles from umbilical cord and postnatal blood with placental histopathology to (1) characterise the intrauterine immune environment of preterm infants compared to term-born controls, and (2) test the hypothesis that exposure to histologic chorioamnionitis is associated with an altered immune and neurotrophic profile in the first week after very preterm birth.

## Materials and Methods

### Study population

Participants were 177 infants, 118 very preterm (mean gestational age 29^+0^ weeks, range 23^+2^ to 32^+0^) and 59 term-born controls, delivered at the Royal Infirmary of Edinburgh, UK and recruited to a longitudinal study of the effect of preterm birth on brain development ^30^. Ethical approval was obtained from the UK National Research Ethics Service and parents provided written informed consent (South East Scotland Research Ethics Committee 16/SS/0154). Subsets were used to characterise the umbilical cord blood immune profile of preterm infants when compared to term-born controls (n=114), and the immune profile on postnatal day 5 in preterm infants who were exposed to HCA compared to those not exposed (n=96).

### Dried blood spot sample analysis

Dried blood spot samples (DBSS) were taken from the umbilical cord following delivery for both preterm cases and term-born controls. For preterm infants, an additional sample was collected on day 5 of life. A customised multiple sandwich immunoassay based on meso-scale technology was used to measure blood spot levels of Interleukin(IL)1-β, IL-2, IL-4, IL-5, IL-6, IL-8, IL-10, IL-12p70, IL-17, IL-18, Monocyte chemotactic protein-1 (MCP-1), Macrophage inflammatory protein-1α (MIP-1α), Macrophage inflammatory protein-1β (MIP-1β), Tumor necrosis factor-α (TNF-α), Tumor necrosis factor-β (TNF-β), Brain-derived neurotropic factor (BDNF), Granulocyte-macrophage colony-stimulating factor (GM-CSF), Interferon-γ (IFN-γ), C-reactive protein (CRP), matrix-metalloproteinase 9 (MMP-9), Regulated upon activation, normal T cell expressed and secreted (RANTES) and Complement components C3, C5a and C9.

Two 3.2 mm disks from the DBSS were punched into each well of Nunc 96-well polystyrene microwell plates (#277143, Thermo Fisher Scientific). 130 µl extraction buffer (PBS containing 1% BSA (Sigma Aldrich #A4503), 0.5% Tween-20 (#8.22184.0500, Merck Millipore), and complete protease inhibitor cocktail (#11836145001, Roche Diagnostics) was added to each well, and the samples were incubated for 1 hour at room temperature on a microwell shaker set at 900 rpm. The extracts were analysed using U-plex plates (Meso-Scale Diagnostics (MSD), Maryland, US) coated with antibodies specific for IL-1β, IL-2, IL-4, IL-5, IL-6, IL-8, IL-12, IL-17, TNF-α, MIP-1β on one plate (#K15067 customized) and BDNF, GM-CSF, IL-10, IL-18, IFN-γ, TNF-β, MCP-1, MIP-1α on another (#K151AC customized) (both MSD). Supplier’s instructions were followed, and extracts were analysed undiluted. A third multiplex analysis was developed in-house applying extracts diluted 1:10 in diluent 7 (#R54BB, MSD) using antibodies specific for C3 (HYB030-07 and HYB030-06, SSI Antibodies, Copenhagen, Denmark), C5a (10604-MM04 and 10604-MM06, Sino Biological, Eschborn, Germany), C9 (R-plex kit #F21XZ, MSD), MMP-9 (BAF911 and MAB911), RANTES (MAB278 and AF278NA) and CRP (BAM17072 and MAB1701) (all R&D Systems, Minneapolis, US) for coating the U-plex plate and for detection, respectively. Coating antibodies (used at 1 μg/mL, except CRP used at 10 ng/mL) were biotinylated (using EZ-Link Sulfo-NHS-LC-Biotin #21327, Thermo Fisher Scientific) in-house (if not already biotinylated at purchase) and detection antibodies were SULFO-tagged (R91AO, MSD), both at a challenge ratio of 20:1. The following calibrators were used: C3: #PSP-109 (Nordic Biosite, Copenhagen, DK), C5a: #10604-HNAE (Sino Biological), C9: #F21XZ (from R-plex kit, MSD), MMP-9: #911-MP, RANTES: #278-RN and CRP: #1707-CR/CF (all from R&D Systems). Calibrators were diluted in diluent 7, detection antibodies (used at 1 μg/mL, except CRP used at 100 ng/mL) were diluted in diluent 3 (#R50AP, MSD). Controls were made in-house from part of the calibrator solution in one batch, aliquoted in portions for each plate and stored at -20°C until use. The samples were prepared on the plates as recommended from the manufacturer and were immediately read on the QuickPlex SQ 120 (MSD). Analyte concentrations were calculated from the calibrator curves on each plate using 4PL logistic regression using the MSD Workbench software. Intra-assay variations were calculated from 16 measurements of a pool of the same control sample on the same plate. Inter-assay variations were calculated from controls analysed in duplicate on each plate during the sample analysis, 4 plates in total. Limits of detection were calculated as 2.5 standard deviations from duplicate measurements of the zero calibrator. The higher detection limit was defined as the highest calibrator concentration. Median intra-assay variation was 8.2% and median inter-assay variation was 11.1%. Detection limits are detailed in Table S1.

### Placental examination

Placental examination was performed by an experienced perinatal pathologist (M.J.E.) and placental reaction patterns were reported according to the site of inflammation, using a structured system ^31^. HCA was defined as the presence of an inflammatory response in the placental membranes of any grade or stage. Maternal inflammatory response (MIR) was defined as the presence of chorionitis, chorioamnionitis or intervillositis. Fetal inflammatory response (FIR) was defined as the presence of vasculitis in the chorionic plate or funisitis involving any vessel of the umbilical cord.

### Statistical analysis

Participant characteristics were compared using Student’s T test or Mann-Whitney U to compare distributions, and Chi-square tests were used to compare proportions. Analytes with values less than the level of detection (<LOD) were assigned the lowest detectable level prior to statistical analysis, and analytes with concentrations <LOD in ≥75% of samples were excluded from subsequent statistical analysis.

To investigate group differences in blood immune mediator profiles, the Mann-Whitney U was used with Bonferroni correction for multiple tests. Principal component analysis (PCA) was used to identify analytes contributing to variance in the cord blood profile and analytes that contributed to PCs with eigenvalues >1 were then entered as independent variables in a logistic regression model to predict preterm or term category. Analytes contributing to variability within PCs predictive of gestational category were then investigated individually using Spearman’s rank order correlation to identify developmentally regulated analytes most strongly correlated with gestational age.

For mediators differentially expressed on day 5, exploratory analyses were performed to characterise the trajectory of immune mediators from birth to postnatal day 5 in association with HCA. Statistical analyses were performed using SPSS version 24.0 (IBM Corp., Armonk, NY), with the exception of PCA, which was performed using R version 3.6.1 (R Core Team, 2019).

## Results

### Participants

The clinical characteristics of participants are shown in Table 1.

**Table 1.** Clinical characteristics of participants.

|  | **Preterm n= 118** | **Term n= 59** |
| --- | --- | --- |
| Mean gestational age, weeks (range) | 29<sup>+0</sup> (23<sup>+2</sup> to 32<sup>+0</sup>) | 39<sup>+4</sup> (37<sup>+3</sup> to 42<sup>+0</sup>) |
| Birthweight, g (SD) | 1213 (408) | 3549 (515) |
| Male sex, n (%) | 67 (57) | 30 (51) |
| Maternal demographics |  |  |
| BMI, mean (SD) | 27.7 (7.3) | 26.4 (5.5) |
| Current smoker, n (%) | 18 (15) | 1 (2) |
| SIMD2016 quintile, n (%) |  |  |
| 1 | 23 (20) | 5 (8) |
| 2 | 22 (19) | 6 (10) |
| 3 | 26 (22) | 11 (19) |
| 4 | 20 (17) | 17 (29) |
| 5 | 23 (20) | 20 (34) |
| Not known | 4 |  |
| Pregnancy complications, n (%) |  |  |
| Pre-eclampsia | 13 (11) | 3 (5) |
| Prolonged rupture of membranes | 27 (23) | 0 (0) |
| Delivery mode, n (%): |  |  |
| Vaginal | 48 (41) | 23 (39) |
| Emergency caesarean | 70 (59) | 10 (17) |
| -Pre-labour | 48 (69) | 1 (10) |
| -In labour | 22 (31) | 9 (90) |
| Elective caesarean | 0 (0) | 26 (44) |
| Any labour, n (%) | 70 (59) | 32 (54) |
| Histologic chorioamnionitis, n (%) | 41 (35) | 10 (17) |
| MIR+ FIR- | 18 (44) | 2 (20) |
| MIR+ FIR+ | 23 (56) | 8 (80) |
| Antenatal steroids (%) | 112 (95) | NA |
| Antenatal magnesium sulphate (%) | 108 (92) | NA |
| Early onset sepsis (%) | 9 (8) | 0 |
| Late onset sepsis (%) | 14 (12) | 0 |
| Bronchopulmonary dysplasia (%) | 32 (27) | NA |
| Necrotizing enterocolitis (%) | 8 (7) | NA |
| Retinopathy of prematurity (%) | 8 (7) | NA |
| Deaths before discharge (%) | 10 (9) | NA |

SIMD2016: Scottish Index of Multiple Deprivation 2016. Prolonged rupture of membranes for >24 hours before delivery. Sepsis: Positive blood culture with a pathogenic organism and/or antibiotic treatment course for ≥5 days. Early-onset sepsis: <72 hours after birth, late-onset sepsis: >72 hours after birth. Bronchopulmonary dysplasia: supplemental oxygen therapy or respiratory support at 36<sup>+0</sup> weeks gestational age. Necrotizing enterocolitis: medical treatment for ≥7 days or surgical treatment. Retinopathy of prematurity requiring treatment with laser therapy. Prolonged rupture of membranes >24 hours.

### Umbilical cord blood profile associated with preterm birth

DBSS were obtained from the umbilical cord of 55 preterm infants and 59 term-born controls. 10 analytes (GM-CSF, IFN-γ, IL-2, IL-4, IL-5, IL-10, IL-12, IL-17, MIP-1α and TNF-β) were <LOD in ≥75% of samples and were therefore excluded from subsequent analysis. Median and interquartile range of analytes are shown in Table 2.

**Table 2.** Cord blood analytes in preterm infants and term-born controls.

|  | Preterm n=55 |  | Term n=59 |  |  |
| --- | --- | --- | --- | --- | --- |
| Analyte (pg/ml) | Median | Q1, Q3 | Median | Q1, Q3 | p-value |
| BDNF | 22.27 | 6.04, 36.90 | 62.33 | 43.18, 113.68 | <0.001 |
| C3 | 3081995.73 | 1858060.85, 4569905.86 | 4447458.83 | 3520957.75, 5613382.51 | <0.001 |
| C5a | 3694.52 | 2420.56, 11470.85 | 7136.48 | 4059.83, 9443.64 | 0.028 |
| C9 | 1236.73 | 215.00, 6901.99 | 12628.69 | 3608.60, 43156.07 | <0.001 |
| CRP | 100.88 | 89.00, 12054.79 | 89.00 | 89.00, 89.00 | <0.001 |
| IL-1β | 0.26 | 0.26, 0.49 | 0.11 | 0.02, 0.34 | 0.009 |
| IL-6 | 0.45 | 0.45, 6.00 | 0.45 | 0.45, 0.45 | <0.001 |
| IL-8 | 12.05 | 5.75, 81.08 | 8.29 | 4.89, 14.71 | 0.049 |
| IL-18 | 25.62 | 10.35, 38.34 | 41.27 | 27.50, 54.78 | <0.001 |
| MCP-1 | 109.09 | 68.69, 210.75 | 48.12 | 38.54, 66.63 | <0.001 |
| MIP-1β | 12.01 | 7.47, 18.82 | 9.72 | 6.88, 18.24 | 0.397 |
| MMP-9 | 14000.68 | 4485.50, 40077.02 | 155885.89 | 87194.67, 358084.73 | <0.001 |
| RANTES | 1460.12 | 702.74, 2754.37 | 3127.55 | 1817.10, 5295.39 | <0.001 |
| TNF-α | 0.27 | 0.27, 0.37 | 0.27 | 0.27, 0.27 | 0.521 |

There were significant group differences for 9 immune mediators (p<0.004, Bonferroni corrected). Pro-inflammatory proteins IL-6, MCP-1 and CRP were elevated in the cord blood of preterm infants whilst BDNF, C3, C9, IL-18, MMP-9 and RANTES were decreased compared to controls born at term. PCA showed that five principal components (eigenvalues>1) explained 76% of the variance in the cord blood profile with the majority of variance explained by the first two components (25% and 20% respectively, Table S2). Projection of individual inflammatory profiles onto the first two principal components is shown in Figure 1.

**Figure 1.**
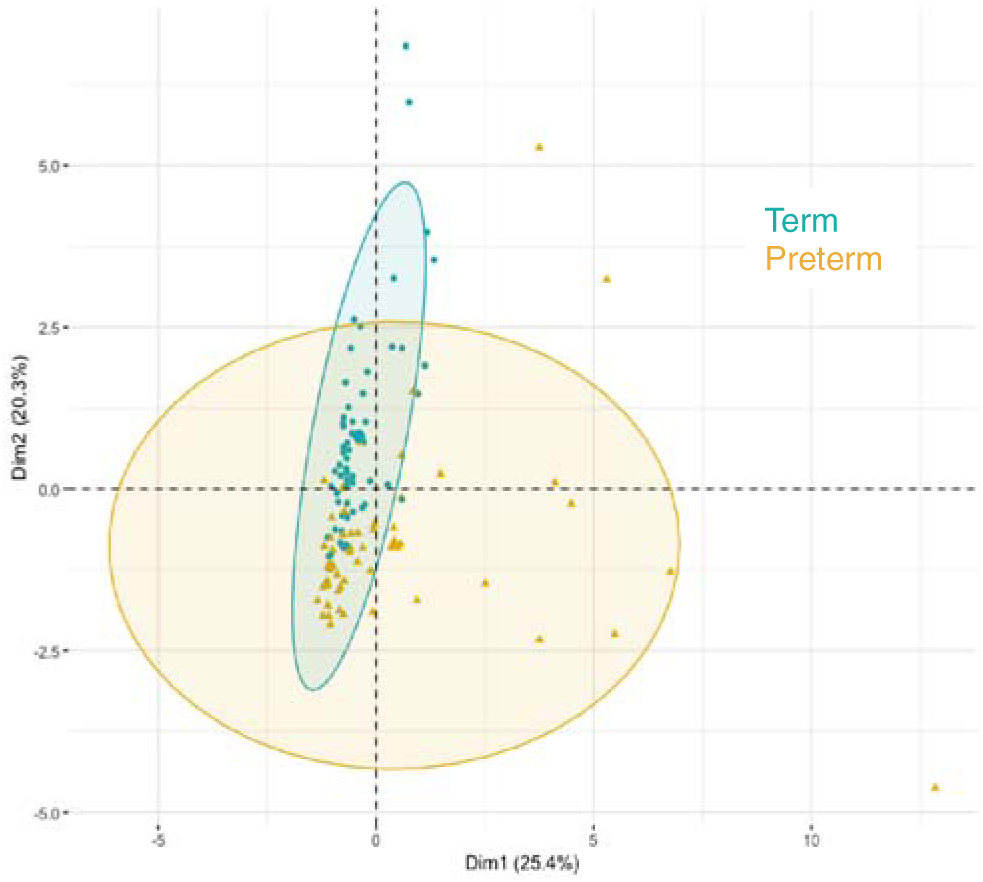
Projection of individual cord blood inflammatory profiles onto the first two principal components, grouped by gestational age category.

In a logistic regression model to predict preterm or term category based on cord blood profile, principal components predicted gestational age category with a classification accuracy of 86% (95% CI 0.78-0.92), p value= 1.242×10^−14^ (Table S3). The percentage contribution of each analyte to the principal components is shown in Figure S1. Amongst immune mediators contributing to variability within the principal components that predicted gestational category, correlation analysis showed that cord blood MMP-9 and BDNF were highly correlated with gestational age at birth (rho >0.65, p<0.001) (Table S4).

### Histologic chorioamnionitis is associated with an altered immune profile on day 5 after very preterm birth

Of 96 preterm infants with day 5 samples, 31 (32%) were exposed to HCA. Infants with HCA exposure had lower GA at birth than infants without HCA: mean GA 28^+2^ weeks versus 29^+4^ weeks (p=0.004). Infants exposed to HCA were more likely to have been delivered vaginally (p<0.001) and more likely to have prolonged rupture of membranes prior to delivery (p<0.001). There were no statistically significant group differences in birthweight, infant sex, exposure to antenatal corticosteroids or magnesium sulphate for fetal neuroprotection, or early onset sepsis (Table S5).

5 immune proteins on day 5 of life had a median level in preterm infants exposed to HCA outside the IQR for preterm infants who were not exposed: BDNF, C3a, IL-8, MIP-1β and MMP-9 (Table S6). In exploratory analyses we considered the trajectory of analytes over time in a subset of 33 preterm infants who had blood obtained from the umbilical cord at delivery and on postnatal day 5. Fourteen infants (42%) were exposed to HCA and eight of those (57%) had evidence of FIR. The 5 mediators that were elevated on day 5 in association with HCA showed markedly different trajectories from birth, and this was influenced by placental histological evidence of a fetal inflammatory response (Figure 2).

**Figure 2.**
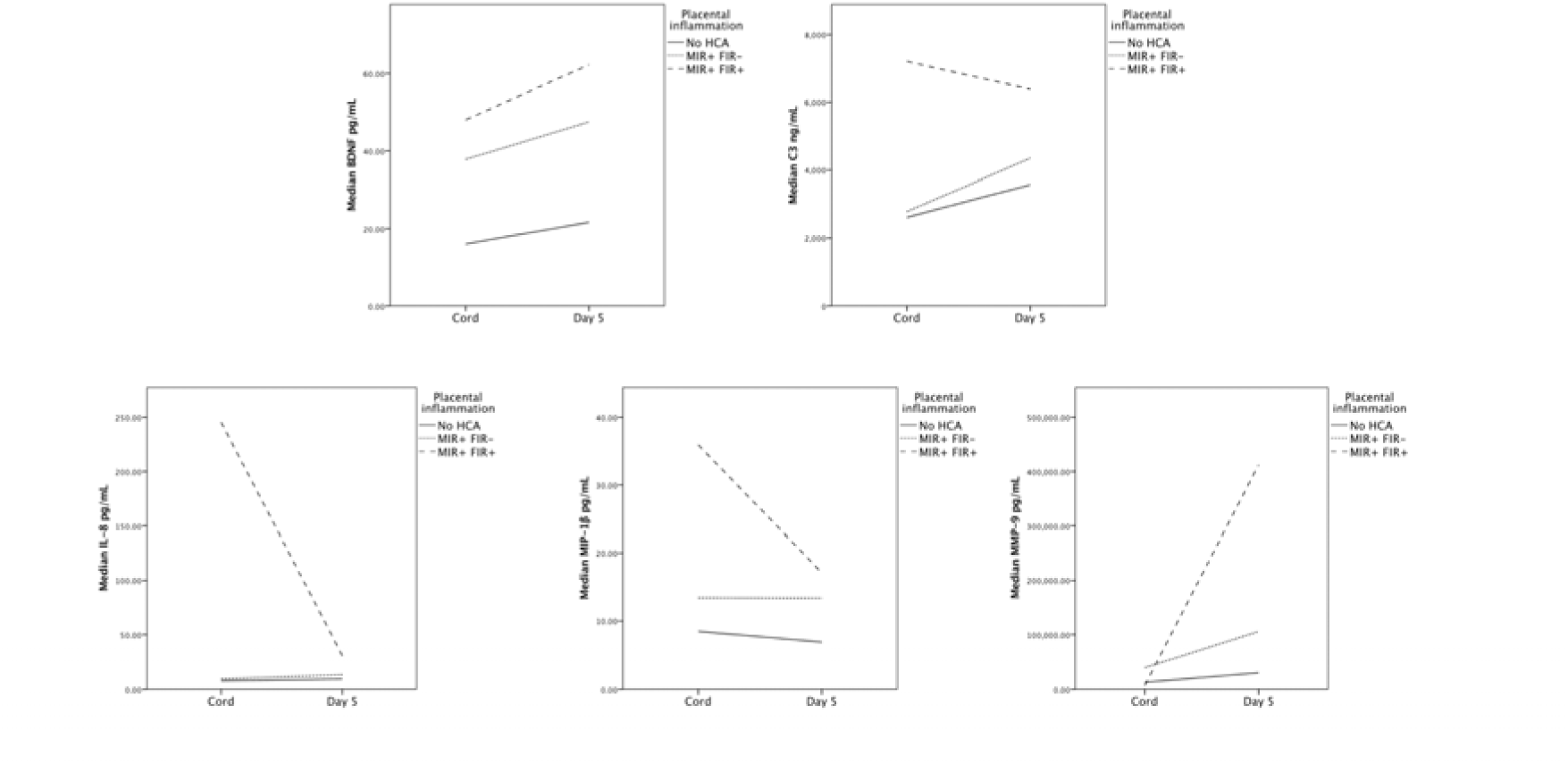
Analyte trajectories from umbilical cord to postnatal day 5 in preterm infants exposed to histologic chorioamnionitis compared to those not exposed.

BDNF and C3 were elevated at both time points in infants with evidence of placental FIR. The differences in cord blood concentrations of IL-8 and MIP-1β in infants with FIR compared to those without were less discriminatory by day 5 whilst altered MMP-9 expression emerged in the postnatal period.

## Discussion

### Main findings

By combining placental histopathology with a customised array of immune mediators in umbilical cord blood and postnatal blood from a large group of mother-infant dyads, this study characterises differences in the systemic immune profile of very preterm infants compared with term-born controls, and demonstrates that exposure to HCA with evidence of a fetal inflammatory response is associated with postnatal immune dysregulation on day 5 after birth.

The umbilical cord blood immune profile was distinctly pro-inflammatory in preterm infants with significant elevations in proteins associated with the acute phase response: IL-6, MCP-1 and CRP. In contrast, six proteins were elevated in healthy term-born controls, suggesting developmental regulation with increasing gestational age: BDNF, C3, C9, IL-18, MMP-9 and RANTES. Five proteins were increased on postnatal day 5 in preterm infants exposed to HCA compared to preterm infants without HCA: BDNF, C3, IL-8, MIP-1β and MMP-9. Our findings are consistent with previous studies showing that the neonatal systemic inflammatory response can be dysregulated and prolonged in the weeks after preterm birth ^4, 27, 28, 32^, but additionally suggest that this is programmed by a fetal inflammatory response.

### Strengths and limitations

We investigated a large number of inflammation-associated proteins representative of the perinatal immune response in the newborn, and used a data driven approach to characterise the inflammatory profile associated with very preterm birth and exposure to HCA.

A limitation of the study is that the concentration of anti-inflammatory cytokines IL-4 and IL-10 were below the level of detection in our participants and so inferences about the balance of damaging and protective factors could not be explored. Another limitation is that amniotic fluid was not available for microbial analysis. Recent transcriptomic studies have shown that the presence of a fetal inflammatory response is more strongly associated with microbial invasion rather than sterile inflammation^33^ but an alternative study design would be required to investigate differences in the postnatal immune profile of infants exposed to intra-amniotic infection compared to sterile inflammation.

Whilst we found an altered trajectory of immune mediators in association with a fetal inflammatory response on postnatal day 5, a larger sample size would be required to perform sub-group analyses based on gestational age or sex and to investigate possible confounding by postnatal events.

### Interpretation

BDNF expression has previously been shown to correlate with gestational age and postnatal age ^29, 34^ but here we show upregulation in preterm infants exposed to HCA. BDNF belongs to the family of neurotrophins: an important group of signaling molecules responsible for neuronal growth, maturation and synaptic plasticity during development ^35^. Prematurity, placental dysfunction and fetal growth restriction have all been associated with reduced levels of BDNF ^29, 36-38^ which may have important implications for long-term brain health. Reduced BDNF in the neonatal period has been associated with increased risk of developing ASD ^39^ whilst elevated BDNF in the weeks after preterm birth is associated with better cognitive performance in childhood ^40, 41^.

C3 was identified as a novel postnatal marker of exposure to intrauterine inflammation. The complement cascade plays a key role in the innate immune response ^42^ but is a potent inflammatory system which when dysregulated can cause significant tissue damage following injury. The complement cascade can be activated through several mechanisms, but all component pathways converge at Complement protein C3 ^43^. C3 participates in multiple key processes affecting developing brain architecture, including tagging of synapses for pruning by microglia ^44, 45^. The complement system is under-developed in preterm infants and complement regulators are low ^46, 47^, which may contribute to an uncontrolled complement response in the context of inflammation ^48^. We have previously shown that the downstream anaphylatoxin, C5a is elevated in the CSF of preterm infants when compared to term-born controls ^6^ and numerous studies beyond the neonatal period have also implicated complement C3 dysregulation in CNS pathology including neurodevelopmental disorders ^49^, multiple sclerosis ^50^, traumatic brain injury ^51^ and neurodegeneration ^52^.

MMP-9 on postnatal day 7 has previously been shown to correlate with the severity of funisitis following extremely preterm birth^27^. MMP-9 is a member of the zinc-dependent endopeptidases that prototypically cleave extracellular matrix (ECM), cell adhesion molecules and cell surface receptors. Matrix-metalloproteinases also modulate the inflammatory response through the regulation of endothelial barrier function, cytokine activity and chemotactic gradient formation ^53^.

The ECM is a key regulator of neural network development and plasticity through the stabilization of synaptic contacts. Dysregulation of MMP-9 during a critical window of CNS vulnerability may therefore have long-term consequences on structural connectivity ^54^. MMP-9 is higher in the CSF of preterm infants when compared to term-born controls and also higher amongst preterm infants with post haemorrhagic ventricular dilatation (PHVD) when compared to those without brain injury ^6,55^. Elevated plasma MMP-9 is also associated with hypoxic-ischemic encephalopathy, correlating with severity of injury in human infants born at term ^56-58^.

Our data suggest that systemic fetal inflammation modulates neurotrophic capability and complement system activation in the perinatal period. Future work is needed to explore whether this altered immune profile extends beyond the first weeks of life and to investigate the relationship between these immune mediators and lung disease, gastrointestinal complications and neurodevelopmental outcomes following preterm birth.

## Conclusions

By combining placental histopathology with a comprehensive assessment of immune mediators, we have shown that very preterm infants have a distinct pro-inflammatory profile in umbilical cord blood and the immune profile of infants exposed to HCA remains altered on day 5 after birth. These results focus research attention on improved detection of fetuses exposed to intrauterine inflammation and they suggest there may be a therapeutic window for targeted intervention that could reduce the risk of co-morbidities associated with HCA.

## Data Availability

Data is available from authors on request.

## Acknowledgements

We are grateful to the families who consented to participate in the study.

## Contribution to authorship

G.S. conceived and designed the study, acquired and analysed data, and drafted the article.

P.G., M.J.E. analysed data and revised the article critically for important intellectual content.

N.B-L., K.S. acquired data, analysed data and revised the article critically for important intellectual content.

D.Q.S., G.J.L. acquired data, and revised the article critically for important intellectual content.

S.C. supervised acquisition of data and revised the article critically for important intellectual content.

J.P.B. conceived and designed the study, supervised acquisition of data, analysed data and drafted the article.

All authors approved the final submitted version.

## Details of ethics approval

Ethical approval was obtained from the UK National Research Ethics Service (South East Scotland Research Ethics Committee 16/SS/0154).

