## Supporting information for "Preterm birth is associated with immune dysregulation which persists in infants exposed to histologic chorioamnionitis: a descriptive study"

**Table S1. Dried blood spot sample detection limits and assay variations.**

| **Analyte** | **Lower detection limit (pg/mL)** | **Higher detection limit (pg/mL)** | **Intra-assay CV%** | **Inter-assay CV%** |
| --- | --- | --- | --- | --- |
| BDNF | 1.05 | 41000 | 2.71 | 3.84 |
| C3 | 13739 | 50000000 | 3.62 | 15.20 |
| C5a | 544 | 16700000 | 5.60 | 19.10 |
| C9 | 8.38 | 3300000 | 5.60 | 12.80 |
| CRP | 89.0 | 100000000 | 16.70 | 34.50 |
| GM-CSF | 0.101 | 12500 | 4.19 | 7.86 |
| IFN-γ | 0.646 | 34875 | 3.24 | 6.29 |
| IL-1β | 0.0260 | 4788 | 11.90 | 10.80 |
| IL-2 | 0.340 | 2688 | 11.30 | 14.70 |
| IL-4 | 0.00956 | 2700 | 16.10 | 16.80 |
| IL-5 | 0.125 | 5125 | 12.00 | 9.67 |
| IL-6 | 0.452 | 2650 | 11.10 | 20.9 |
| IL-8 | 0.0684 | 2575 | 10.9 | 15.0 |
| IL-10 | 0.0886 | 4625 | 4.13 | 5.56 |
| IL-12p70 | 0.157 | 9063 | 10.30 | 17.00 |
| IL-17 | 0.286 | 47375 | 13.50 | 11.40 |
| IL-18 | 0.199 | 50500 | 3.36 | 6.58 |
| MCP-1 | 1.28 | 7400 | 2.55 | 6.99 |
| MIP-1α | 0.960 | 7500 | 2.71 | 6.32 |
| MIP-1β | 1.73 | 2325 | 13.8 | 5.39 |
| MMP-9 | 24.1 | 5000000 | 6.29 | 25.10 |
| RANTES | 37.9 | 1600000 | 10.10 | 29.00 |
| TNF-α | 0.272 | 4538 | 11.80 | 4.48 |
| TNF-β | 0.0250 | 5050 | 2.52 | 3.90 |

**Table S2. Variance in the cord blood inflammatory profile.**

| **Component** | **Eigenvalue** | **Total**  **variance /%** | **Cumulative variance /%** |
| --- | --- | --- | --- |
| 1 | 3.56 | 25.43 | 25.43 |
| 2 | 2.85 | 20.33 | 45.76 |
| 3 | 1.88 | 13.44 | 59.20 |
| 4 | 1.29 | 9.21 | 68.42 |
| 5 | 1.10 | 7.84 | 76.25 |
| 6 | 0.85 | 6.07 | 82.32 |
| 7 | 0.58 | 4.13 | 86.45 |
| 8 | 0.50 | 3.55 | 90.00 |
| 9 | 0.46 | 3.32 | 93.31 |
| 10 | 0.31 | 2.22 | 95.53 |
| 11 | 0.20 | 1.45 | 96.98 |
| 12 | 0.19 | 1.38 | 98.36 |
| 13 | 0.18 | 1.31 | 99.67 |
| 14 | 0.05 | 0.33 | 100.00 |

**Table S3. Logistic regression for the prediction of gestational age category using principal components derived from the umbilical cord blood profile.**

|  | **B** | **β** | **p-value** |
| --- | --- | --- | --- |
| PC1 | 2.8189 | 5.3424 | 0.000337 |
| PC2 | -2.6162 | -4.4331 | 1.71e-06 |
| PC3 | -0.1343 | -0.1850 | 0.648519 |
| PC4 | 0.8840 | 1.0084 | 0.135172 |
| PC5 | -0.8628 | -0.9076 | 0.027207 |

**Table S4. Correlation between individual analytes that contributed to the principal components predictive of gestational category and gestational age at birth.**

| **Analyte** | **Spearman’s rho** | **p-value** |
| --- | --- | --- |
| MMP-9 | 0.685 | 4.1162x10^-17^ |
| BDNF | 0.654 | 2.8795x10^-15^ |
| RANTES | 0.346 | 0.000160 |
| C3 | 0.290 | 0.002 |
| IL-1β | 0.190 | 0.043 |

**Table S****5. Characteristics of preterm infants with day 5 samples who were exposed to histologic chorioamnionitis compared to those not exposed.**

|  | **No HCA n= 65** | **HCA n= 31** |
| --- | --- | --- |
| Gestational age, weeks (SD) | 29^+4^ (1.84) | 28^+2^ (2.63) |
| Birthweight, g (SD) | 1246 (371) | 1187 (421) |
| Male sex, n (%) | 37 (57) | 18 (58) |
| Delivery mode, n (%):  Vaginal  Caesarean  Pre labour  In labour  Any labour, n (%) | 15 (23)  50 (77)  34 (68)  16 (32)  31 (48) | 23 (74)  8 (26)  8 (100)  0 (0)  23 (74) |
| Antenatal steroids, n (%) | 61 (94) | 30 (97) |
| Magnesium sulphate, n (%) | 59 (91) | 30 (97) |
| Prolonged rupture of membranes, n (%) | 8 (12) | 14 (45) |
| Early onset sepsis, n (%) | 4 (6) | 4 (13) |

Prolonged rupture of membranes for >24 hours before delivery. Sepsis: Positive blood culture with a pathogenic organism and/or antibiotic treatment course for ≥ 5 days. Early-onset sepsis: <72 hours after birth.

**Table S6. Day 5 blood analyte concentrations for preterm infants exposed to histologic chorioamnionitis compared to those not exposed.**

|  | **No HCA n=65** | | **HCA n=31** | |  |
| --- | --- | --- | --- | --- | --- |
| **Analyte (pg/ml)** | **Median** | **Q1,Q3** | **Median** | **Q1,Q3** | **p-value** |
| **BDNF** | **28.49** | **14.27, 42.74** | **42.28** | **26.16, 100.09** | **0.008** |
| **C3** | **3584453.76** | **2761102.54, 4875046.42** | **5084306.96** | **4180179.50, 6912830.38** | **<0.001** |
| C5a | 6457.25 | 4218.14, 9987.78 | 9504.83 | 7336.94, 13450.98 | 0.002 |
| C9 | 7462.57 | 2455.08, 20710.16 | 18110.18 | 4761.32, 55877.40 | 0.028 |
| CRP | 248.98 | 89.00, 8104.18 | 1431.89 | 89.00, 25859.42 | 0.194 |
| IL-1β | 0.03 | 0.03, 0.08 | 0.05 | 0.03, 0.11 | 0.065 |
| IL-6 | 0.45 | 0.45, 0.45 | 0.45 | 0.45, 0.45 | 0.641 |
| **IL-8** | **11.44** | **7.61, 19.30** | **29.26** | **12.55, 39.53** | **0.012** |
| IL-18 | 35.78 | 21.10, 53.64 | 34.35 | 21.98, 47.92 | 0.501 |
| MCP-1 | 135.02 | 108.41, 212.56 | 106.97 | 90.22, 148.65 | 0.010 |
| **MIP-1β** | **7.88** | **5.50, 10.95** | **12.04** | **7.22, 16.22** | **0.011** |
| **MMP-9** | **56654.38** | **28317.00, 99300.57** | **240084.32** | **64198.64, 460436.84** | **<0.001** |
| RANTES | 2606.92 | 910.54, 4199.97 | 3486.66 | 2058.30, 5362.46 | 0.070 |
| TNF-α | 0.27 | 0.27, 0.27 | 0.27 | 0.27, 0.27 | 0.986 |

Analytes with a median level in preterm infants exposed to HCA outside the IQR for preterm infants without HCA are highlighted in bold.

**Figure S1. The percentage contribution of each analyte to variability in the cord blood profile.**

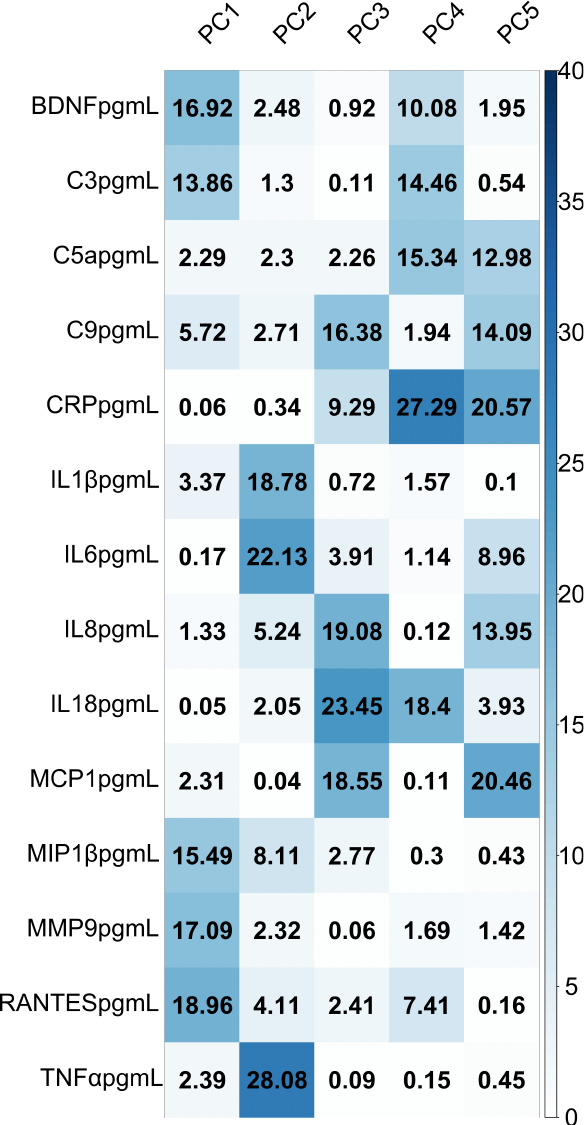
